# Variation in False Negative Rate of RT-PCR Based SARS-CoV-2 Tests by Time Since Exposure

**DOI:** 10.1101/2020.04.07.20051474

**Authors:** Lauren M Kucirka, Stephen A Lauer, Oliver Laeyendecker, Denali Boon, Justin Lessler

## Abstract

SARS-CoV-2 RT-PCR based tests are being used to “rule out” infection among high-risk individuals such as exposed inpatients and healthcare workers. It is critical to understand how the predictive value of the test varies with time from exposure and symptom onset in order to avoid being falsely reassured by negative tests. As such, the goal of our study was to estimate the false negative rate by day since infection. We used previously published data on RT-PCR sensitivity on samples derived from nasal swabs by day since symptom onset (n=633) and fit a cubic polynomial spline to calculate the false negative rate by day since exposure and symptom onset. Over the four days of infection prior to the typical time of symptom onset (day 5) the probability of a false negative test in an infected individual falls from 100% on day one (95% CI 69-100%) to 61% on day four (95% CI 18-98%), though there is considerable uncertainty in these numbers. On the day of symptom onset, the median false negative rate was 39% (95% CI 16-77%). This decreased to 26% (95% CI 18-34%) on day 8 (3 days after symptom onset), then began to rise again, from 27% (95% CI 20-34%) on day 9 to 61% (95% CI 54-67%) on day 21. Care must be taken when interpreting RT-PCR tests for SARS-CoV-2 infection, particularly if performed early in the course of infection, when using these results as a basis for removing precautions intended to prevent onward transmission. If there is high clinical suspicion, patients should not be ruled out on the basis of RT-PCR alone, and the clinical and epidemiologic situation should be carefully considered.

## BACKGROUND

SARS-CoV-2 RT-PCR based tests are often used to “rule out” infection among high-risk individuals such as exposed inpatients and healthcare workers. Hence, it is critical to understand how the predictive value changes in relation to time since exposure or symptoms, especially when using the results of these tests to make decisions about whether to stop using personal protective equipment (PPE) or allow exposed healthcare workers to return to work. The sensitivity and specificity of SARS-CoV-2 PCR based tests are poorly characterized and the “window period” after acquisition in which testing is most likely to produce false negative results, is not well known.

Accurate testing for SARS-CoV-2, followed by appropriate preventive measures, is paramount in the healthcare setting to prevent both nosocomial and community transmission. However, most hospitals are facing critical shortages of SARS-CoV-2 testing capacity, PPE, and healthcare personnel (1). As the epidemic progresses, hospitals are increasingly faced with how to respond when a patient or healthcare worker has a known exposure to SARS-CoV-2. While 14 days of airborne precautions or quarantine would be a conservative approach to minimizing transmission per CDC guidelines (2), this is not feasible for many hospitals given starkly limited resources.

As RT-PCR-based tests for SARS-CoV-2 are becoming more available, these are increasingly being used to “rule out” infection to conserve scarce PPE and preserve the workforce. When an exposed healthcare worker tests negative they may be cleared to return to work; similarly when an exposed patient tests negative, airborne/droplet precautions may be removed. If negative tests performed during the window period are treated as strong evidence that an exposed individual is negative, preventable transmission could occur.

## OBJECTIVE

It is critical to understand how the predictive value of the test varies with time from exposure and symptom onset in order to avoid being falsely reassured by negative tests performed early in the course of infection. As such, the goal of our study was to estimate the false negative rate by day since infection.

## METHODS AND FINDINGS

We used previously published data on RT-PCR sensitivity on samples derived from nasal swabs by day since symptom onset for patients in Shenzhen (n=414) and Wuhan (n=219) (4,5). Using an approach similar to Leisenring et al. (1997) and Azman et al. (2020, preprint) (6),(7), we fit a cubic polynomial spline to test sensitivity by log-time since exposure, assumed to have occurred 5 days prior to symptom onset based on the median incubation period (8). We restrict the function to be monotonically increasing for the first four days post exposure (one day prior to symptom onset), and calculate the expected false negative rate on each day. We calculate the post test probability of infection, assuming a prior probability based on the attack rate in close household contacts of SARS-CoV-2 cases in Shenzhen (77/686, 11.2%) (9). We assume a specificity of 100% for RT-PCR.

### Probability of a False Negative Result Among SARS-CoV-2 Positive Patients, by Day Since Exposure

Over the four days of infection prior to the typical time of symptom onset (day 5) the probability of a false negative test in an infected individual falls from 100% on day one (95% CI 69-100%) to 61% on day four (95% CI 18-98%), though there is considerable uncertainty in these numbers.

### Probability of a False Negative Result by Time Since Symptom Onset

On the day of symptom onset, the median false negative rate was 39% (95% CI 16-77%) (Figure 1, upper panel). This decreased to 26% (95% CI 18-34%) on day 8 (3 days after symptom onset), then began to rise again, from 27% (95% CI 20-34%) on day 9 to 61% (95% CI 54-67%) on day 21.

**Figure 1:**
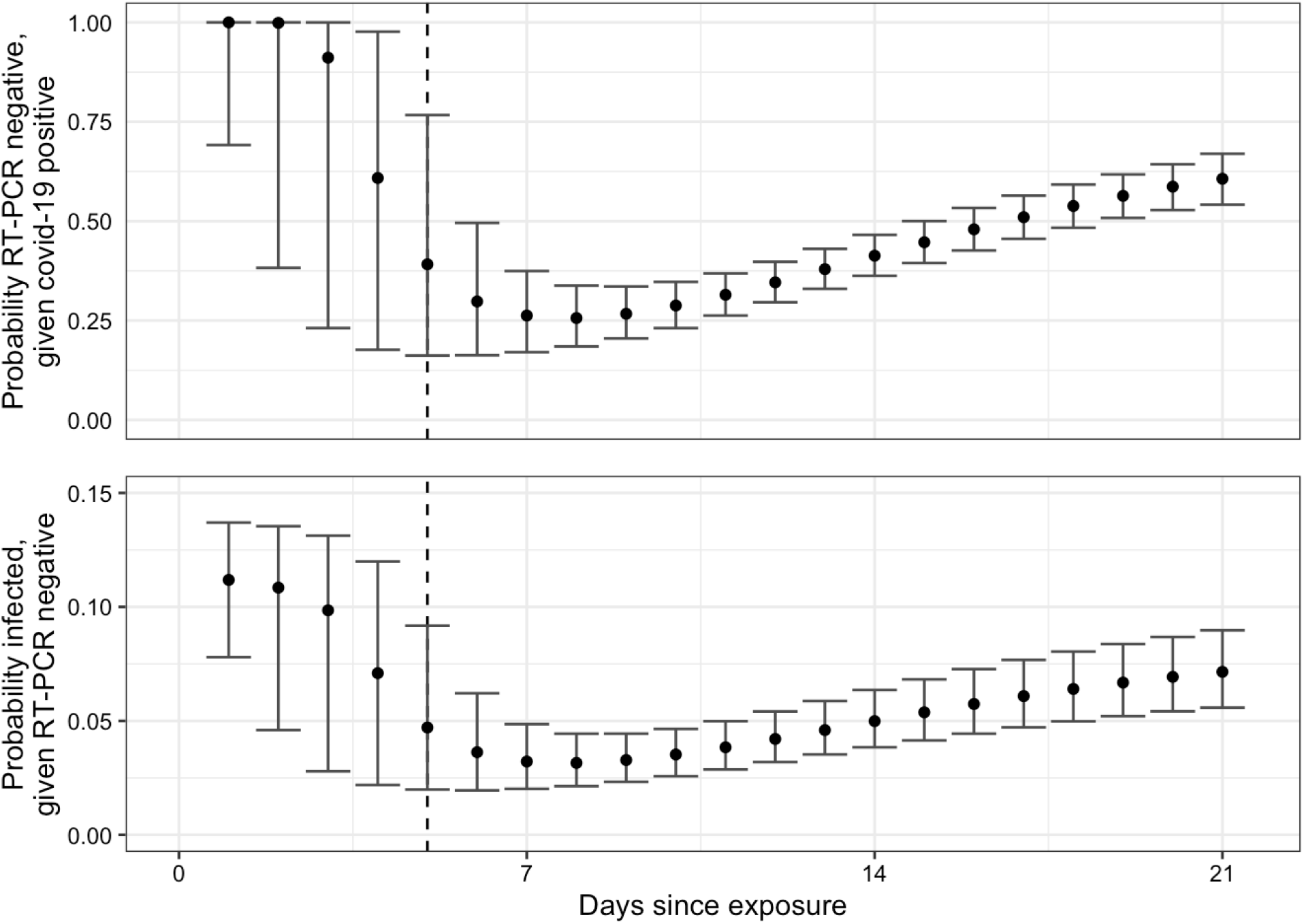
Probability of 1) being RT-PCR positive among SARS-CoV-2 infected patients (Upper Panel), and 2) being infected, given RT-PCR negative (Lower Panel), by days since exposure

### Probability of Infection if RT-PCR Test is Negative

Translating these results into a post test probability of infection, a negative test on day 3 would reduce our estimate of the probability a case was infected by only 8% (95% CI: 0-75%) (e.g., from 11% (95% CI 9-14%) to 10% (95% CI 3-13%) in close household contacts). Tests performed on the first day of symptom onset are more informative, reducing the inferred probability that a case was infected by 58% (95% CI 21-82%).

## DISCUSSION

Care must be taken when interpreting RT-PCR tests for SARS-CoV-2 infection, particularly if performed early in the course of infection, when using these results as a basis for removing precautions intended to prevent onward transmission. If there is high clinical suspicion, patients should not be ruled out on the basis of RT-PCR alone, and the clinical and epidemiologic situation should be carefully considered. In many cases, time of exposure is unknown and testing is performed based on time of symptom onset. The false negative rate is lowest 3 days after onset of symptoms, or approximately 8 days post-exposure. If possible, sputum samples should be tested as they provide higher levels of detectable virus than nasal swabs. This information should be considered when designing testing protocols: e.g. requiring two negative tests before considering a patient non-infected. If limited resources preclude multiple tests, clinicians should consider waiting 2-3 days after symptom onset to minimize the probability of a false negative result. Further studies to characterize test performance and research into higher sensitivity approaches are critical.

## Data Availability

All data from cited publications or publicly available sources

